# Body mass index modifies symptom-specific metabolomic associations with depressive symptoms in the Estonian Biobank

**DOI:** 10.64898/2026.09.01.26361909

**Authors:** Siim Kurvits, Nele Taba, Estonian Biobank research team, Lili Milani, Toomas Haller, Kelli Lehto

## Abstract

**Background:** Metabolomic studies of depression have yielded heterogeneous findings, potentially because metabolic correlates differ across symptoms and metabolic states. We examined symptom-specific metabolomic associations and whether body mass index (BMI) modifies these relationships.

**Methods:** We analyzed 83,717 Estonian Biobank participants (70.6% female) with 249 Nightingale metabolite measures and 14 lifetime depressive symptoms. Logistic regression models progressively adjusted for sociodemographic, lifestyle, medication, and BMI factors. BMI-related attenuation and metabolite × BMI interactions were evaluated, followed by self-organizing map analyses of broader metabolic context.

**Results:** Before BMI adjustment, 660 metabolite–symptom associations were Bonferroni-significant; 136 were significant after BMI adjustment, including 105 retained associations. Weight-related associations showed the strongest BMI dependence: none of 199 weight-gain associations and 2 of 115 weight-loss associations were retained. Among 691 preselected metabolite–symptom pairs, 211 (30.5%) showed significant metabolite × BMI interactions after false discovery rate correction. Six systemic metabolic profiles were identified, but only 3 of 211 BMI-sensitive pairs showed additional profile-dependent heterogeneity.

**Conclusions:** Circulating metabolic correlates of depressive symptoms are heterogeneous and strongly dependent on symptom phenotype and BMI-related metabolic context. These findings suggest that metabolic biomarkers in depression should be interpreted in relation to both symptom presentation and metabolic state rather than as uniform correlates of the disorder.

## Introduction

Major depressive disorder (MDD) is a leading cause of disability and is characterized by substantial clinical and biological heterogeneity (1,2). Individuals meeting diagnostic criteria may present with opposing symptoms, such as insomnia versus hypersomnia or weight loss versus weight gain, yet are grouped within the same diagnostic category (3,4). This heterogeneity challenges the assumption that depression has a uniform biological signature and may obscure symptom-specific mechanisms in conventional case–control studies (5,6). Accordingly, symptom-level approaches have gained prominence as a means of mapping biological signals more directly onto individual depressive features (7,8).

Large-scale metabolomic studies have implicated lipid metabolism, inflammation, and amino acid pathways in depression (9–11), but findings remain inconsistent across studies and populations (12,13). One explanation is that metabolic correlates differ across depressive symptoms and depend on biological context that is rarely modelled explicitly (14). Prior symptom-level metabolomic studies support heterogeneity across symptom domains, but the extent to which body mass index (BMI)-related physiology accounts for or modifies individual metabolite–symptom associations remains unclear (11,14).

Adiposity is strongly associated with cardiometabolic risk and is also linked to common mental disorders (15,16). In psychiatric metabolomic studies, BMI is commonly treated primarily as an adjustment variable (9,10). However, adiposity-linked inflammation and metabolic dysregulation may define biologically meaningful heterogeneity within depression and modify how peripheral metabolic processes relate to depressive symptoms (14,17–19). Whether such BMI-dependent metabolic sensitivity is concentrated in neurovegetative symptoms or also extends to core affective and cognitive symptoms remains insufficiently understood. More broadly, biologically informed stratification has been proposed as an important route toward precision psychiatry (20). Experimental and treatment studies further suggest that inflammatory state may identify clinically relevant heterogeneity within depression: baseline inflammation modified response to tumor necrosis factor antagonism in treatment-resistant depression (21), while experimentally induced inflammation produced greater anhedonic responses among participants with higher baseline C-reactive protein concentrations (22).

Existing large-scale metabolomic studies have substantially advanced understanding of depression-associated metabolic signatures but have predominantly relied on diagnostic case–control designs and have generally treated BMI as an adjustment factor rather than an analytical target (9,10). The Nightingale NMR platform enables population-scale characterization of circulating metabolic traits across national biobanks (23). Here, we examined associations between 14 lifetime depressive symptoms and 249 circulating Nightingale NMR metabolites in 83,717 Estonian Biobank (EstBB) participants with detailed symptom-level assessment (24,25). We quantified changes in metabolite–symptom associations after BMI adjustment and formally tested BMI as an effect modifier. We hypothesized that BMI-related metabolic context would most strongly condition associations with weight- and appetite-related symptoms while also modifying selected affective and cognitive associations. Finally, we derived systemic metabolic profiles in the full population independently of psychiatric phenotypes and tested whether broader metabolic state explained additional heterogeneity beyond continuous BMI. Together, these analyses test whether peripheral metabolic correlates of depression are better understood as symptom- and context-dependent signals rather than as uniform biomarkers of the disorder.

## Methods and Materials

### Study population

The EstBB is a volunteer-based biobank covering approximately 212,000 adults (∼20% of the Estonian adult population) (25). Biobank data are annually linked to national electronic health records covering medical diagnoses and medication prescriptions from primary and specialist care and supplemented by questionnaire-based assessments of lifestyle and mental health.

The present study included participants who completed the 2021 Mental Health Online Survey (MHoS) and had available NMR metabolomic data (N = 83,717) (24, 25). The prevalence of EHR-recorded major depressive disorder among MHoS participants (24.3%) was comparable to non-participating EstBB donors (23.6%) (24).

### Ethics approval and consent

The activities of the EstBB are regulated by the Human Genetic Research Act adopted specifically for the operations of the EstBB. Participants included in this study provided written informed consent at recruitment for the use of their samples and health and genomic data for research. Individual level data analysis in the EstBB was carried out under ethical approval [1.1-12/4561] from the Estonian Committee on Bioethics and Human Research (Estonian Ministry of Social Affairs), using data according to release application [6-7/GI/10083 V16] from the Estonian Biobank.

### Lifetime depression symptoms and BMI

Lifetime depression symptoms were assessed using the depression module of the Composite International Diagnostic Interview - Short Form (CIDI-SF) (24,26). Depressed mood and anhedonia lasting ≥2 weeks were assessed as core symptoms. Secondary depressive symptoms, including changes in weight, appetite and sleep, were presented only to participants endorsing at least one core symptom, according to the questionnaire skip logic. Participants endorsing a secondary symptom were classified as cases for that symptom; participants who did not endorse the secondary symptom, as well as those who did not endorse either core symptom and therefore were not presented with the secondary-symptom questions, were classified as controls. Thus, all symptom analyses were conducted in the full eligible study cohort rather than being restricted to participants with core depressive symptoms.

Height and weight were self-reported in the MHoS and used to calculate BMI (kg/m²). BMI was used as an imperfect proxy for overall adiposity, acknowledging that it does not capture body composition or fat distribution. In sequential models, BMI adjustment was used descriptively to quantify dependency of metabolite–symptom associations on adiposity-linked metabolic variation and should not be interpreted as isolating causal effects.

### Metabolic Biomarkers

Blood plasma samples were obtained from EstBB participants at recruitment and circulating metabolomic profiles were quantified using a high-throughput proton nuclear magnetic resonance (¹H-NMR) spectroscopy platform (Nightingale Health Ltd., Helsinki, Finland). The platform provides standardized quantification of 249 lipid, lipoprotein, fatty acid, and small-molecule metabolite ratios and sums (23).

The recruitment of EstBB was conducted in two waves (ca 25% and 75% of samples) with different recruiting strategies (25) and different sample collection protocols. Time between blood sampling and MHoS completion (deltaT) was included as a covariate in all regression models to account for variation in temporal distance between metabolite measurement and symptom reporting.

### Quality Control and Normalization

Metabolomic quality control followed established recommendations for population-scale NMR data (27). Duplicate samples, withdrawn participants, and samples with >200 missing metabolites were excluded. Technical variation was normalized using remove_technical_variation() from the ukbnmr R package version 2.2. Normalization performance was evaluated across spectrometers and batches. Because normalization introduced batch effects in extremely right-skewed XXL-VLDL measures, these variables were restored to their pre-normalization values. Composite biomarkers were subsequently recomputed and missing ratio values restored.

Metabolites were standardized using sex-specific z-scores. Values with |z| ≥5 were excluded as extreme outliers. Regression analyses used complete cases, whereas missing values were imputed with metabolite-specific means for SOM training.

### Statistical analysis

Continuous variables are presented as mean ± SD, categorical variables as counts and percentages. Group differences were assessed using t-tests, Mann–Whitney U tests, or χ² tests as appropriate. All analyses were conducted in R version 4.3.1.

### Metabolome-wide association Study (MetWAS)

Participants with >10 missing metabolite measurements were excluded from MetWAS analyses. Each depression symptom was modeled as a binary outcome, and metabolites as standardized continuous predictors.

Logistic regression models were fitted using sequential adjustment:

- Model 1 (minimally adjusted): age, sex, deltaT
- Model 2 (lifestyle-adjusted): Model 1 + smoking status, alcohol consumption, education, cardiometabolic medications (ATC A10, C02, C10)
- Model 3 (BMI-adjusted): Model 2 + BMI

Model 2 covariates were selected to account for lifestyle, socioeconomic, and cardiometabolic factors associated with both metabolism and depression. Antidepressant medication was not included because symptoms were assessed retrospectively without temporal anchoring to treatment exposure. Bonferroni correction was applied across 3,486 tests (249 metabolites × 14 symptoms; p < 1.43 × 10□□).

### Quantifying BMI-related attenuation of metabolite–symptom associations

To determine how coefficients changed after adding BMI, we quantified attenuation of effect sizes following BMI adjustment. Attenuation was calculated on the log-odds scale to preserve linearity of effect estimates and avoid distortion inherent to odds ratio–based comparisons. For metabolite–symptom pairs significant in Model 2, BMI-related attenuation was calculated on the log-odds scale as 1-I *β*_3_I/I *β*_2_I, where *β*_2_and *β*_3_are coefficients from Models 2 and 3, respectively. Values approaching 1 indicate strong attenuation, values near 0 little change, and negative values increased effect magnitude after BMI adjustment. Attenuation analyses were descriptive and were not interpreted as distinguishing confounding from mediation.

### BMI as an effect modifier of metabolite–symptom associations

Interaction analyses were restricted to the 691 unique metabolite–symptom pairs reaching Bonferroni significance in Model 2 or Model 3. For each pair, we fitted a logistic regression model including standardized metabolite, standardized BMI, their multiplicative interaction, and Model 2 covariates. Effect modification was assessed using Wald tests of the metabolite × BMI coefficient. Benjamini–Hochberg FDR correction was applied across all 691 tests. Positive interaction coefficients indicate that the metabolite–symptom slope becomes more positive with increasing BMI, whereas negative coefficients indicate that it becomes more negative. Predictions were estimated at BMI −1 SD, mean, and +1 SD for visualization.

### Self-Organizing Map (SOM) analysis

To characterize broader metabolic heterogeneity, we trained a SOM using only the 249 sex-standardized metabolite measures, with metabolite-specific mean imputation for missing values. The final model used a 21 × 21 hexagonal lattice and 20,000 training iterations using the kohonen package (28–30). Detailed training parameters and model diagnostics are provided in Supplementary Note 1.

SOM codebook vectors were grouped using Ward.D2 hierarchical clustering with Euclidean distance. Candidate cluster solutions were evaluated using C-index, Calinski–Harabasz, Davies–Bouldin, Dunn, silhouette, and Wemmert–Gancarski indices. Six broader metabolic profiles were selected based on cluster validation, population coverage, and biological interpretability.

Because local topology preservation was limited (topographic error = 0.81), downstream inference relied on profile membership rather than local adjacency of SOM units.

### Metabolite × SOM-profile interaction analysis

To test whether metabolite–symptom associations showed heterogeneity across systemic metabolic states beyond that captured by continuous BMI, we evaluated metabolite × SOM-profile interactions for the 211 metabolite–symptom pairs showing FDR-significant metabolite × BMI interactions in the preceding analysis. For each pair, a base logistic regression model included the metabolite, BMI, metabolite × BMI interaction, SOM profile, and the same Model 2 covariates. This was compared using a likelihood ratio test with a model additionally containing the metabolite × SOM-profile interaction. P-values were corrected across all 211 tests using the Benjamini–Hochberg FDR procedure. Profile-specific odds ratios were derived from the full model at mean BMI.

## Results

### Study Population and depressive symptom prevalence

A total of 83,717 individuals completed the Mental Health Online Survey and had NMR metabolomic data available (Table 1). Mean age at survey completion was 48 years, and 70.6% were female. In total, 42,938 (51.3%) endorsed at least one of the two core lifetime symptoms (depressed mood and/or anhedonia). Women reported depressive symptoms more frequently than men (55.6% vs 41.0%). Participants with symptoms were slightly younger (birth year 1975 vs 1971) and more likely to smoke, while BMI difference was modest (∼1kg/m^2^). Cardiometabolic medication use was slightly lower among those reporting depression symptoms (Table 1). Depression symptom prevalence ranged from 49.6% (depressed mood) and 42.7% (fatigue) to 6.6% (hypersomnia) and 16.3% (thoughts of death) (Supplementary Table S1).

**Table 1.**
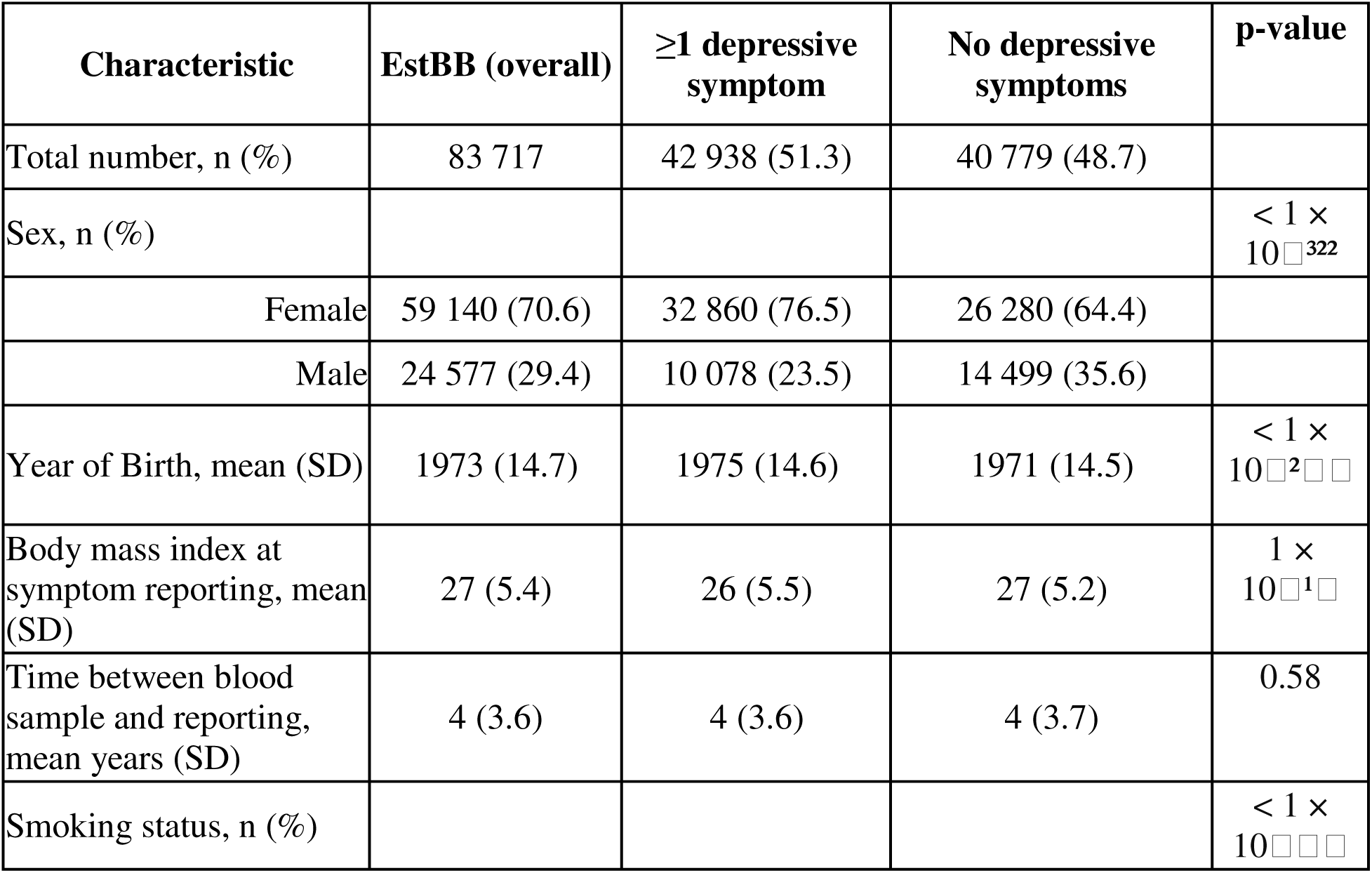
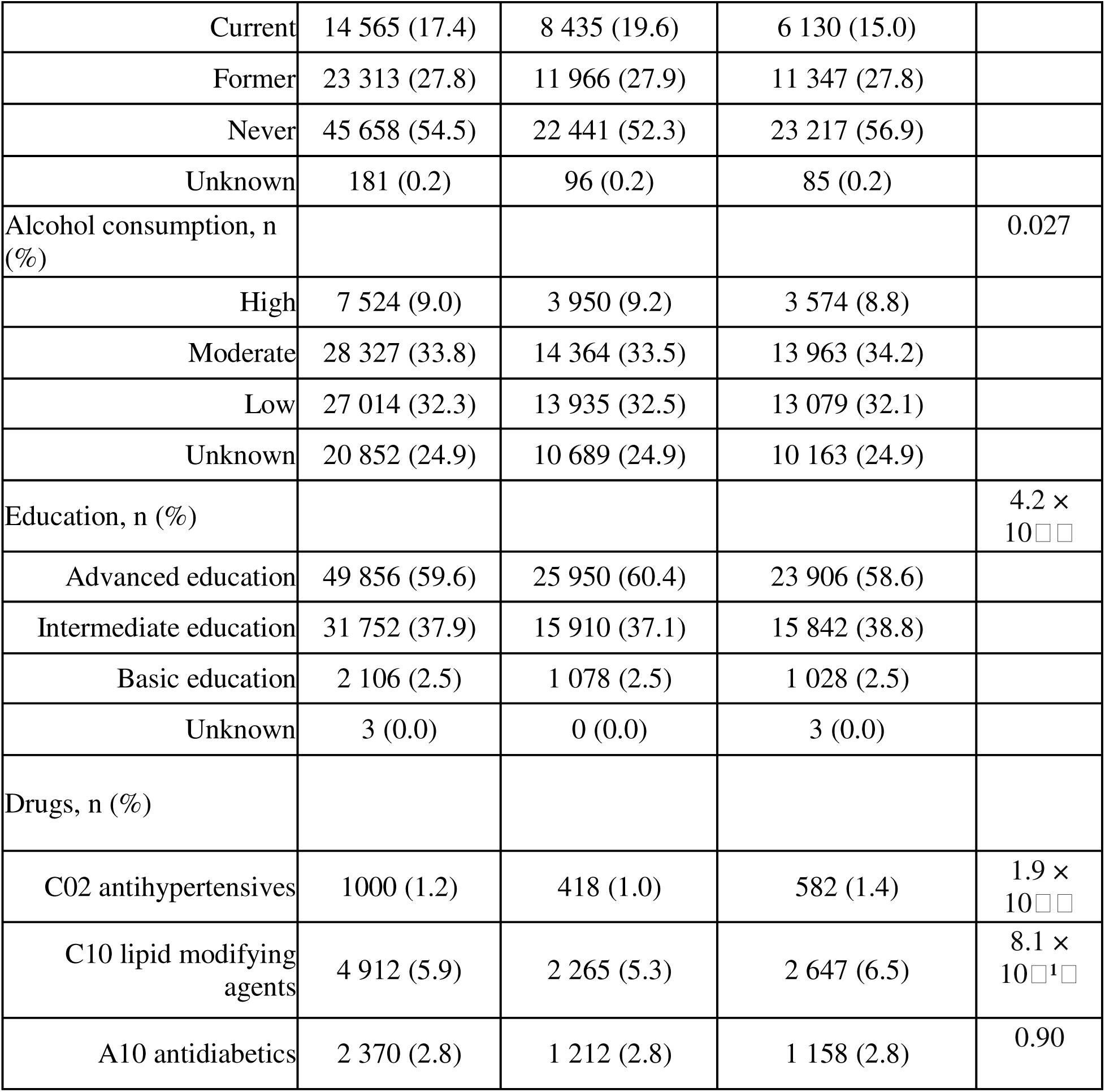
Characteristics of Study Population.

### Symptom-level metabolome-wide association analyses

We tested associations between 249 metabolites and 14 depressive symptoms. Significant associations decreased from 1,367 in Model 1 to 660 after lifestyle and medication adjustment (Model 2) and 136 after additional BMI adjustment (Model 3; Figure 1; Supplementary Tables S2–S4). Associations were concentrated among weight- and appetite-related symptoms, particularly weight gain (199 associations in Model 2) and weight loss (115). These symptoms were broadly associated with triglyceride-rich lipoproteins, GlycA, and fatty-acid composition. For example, weight gain was associated with GlycA (OR 1.27, 95% CI 1.24–1.30) and L-HDL-PL% (OR 1.25, 95% CI 1.22–1.28). Affective and cognitive symptoms generally showed fewer associations.

**Figure 1.**
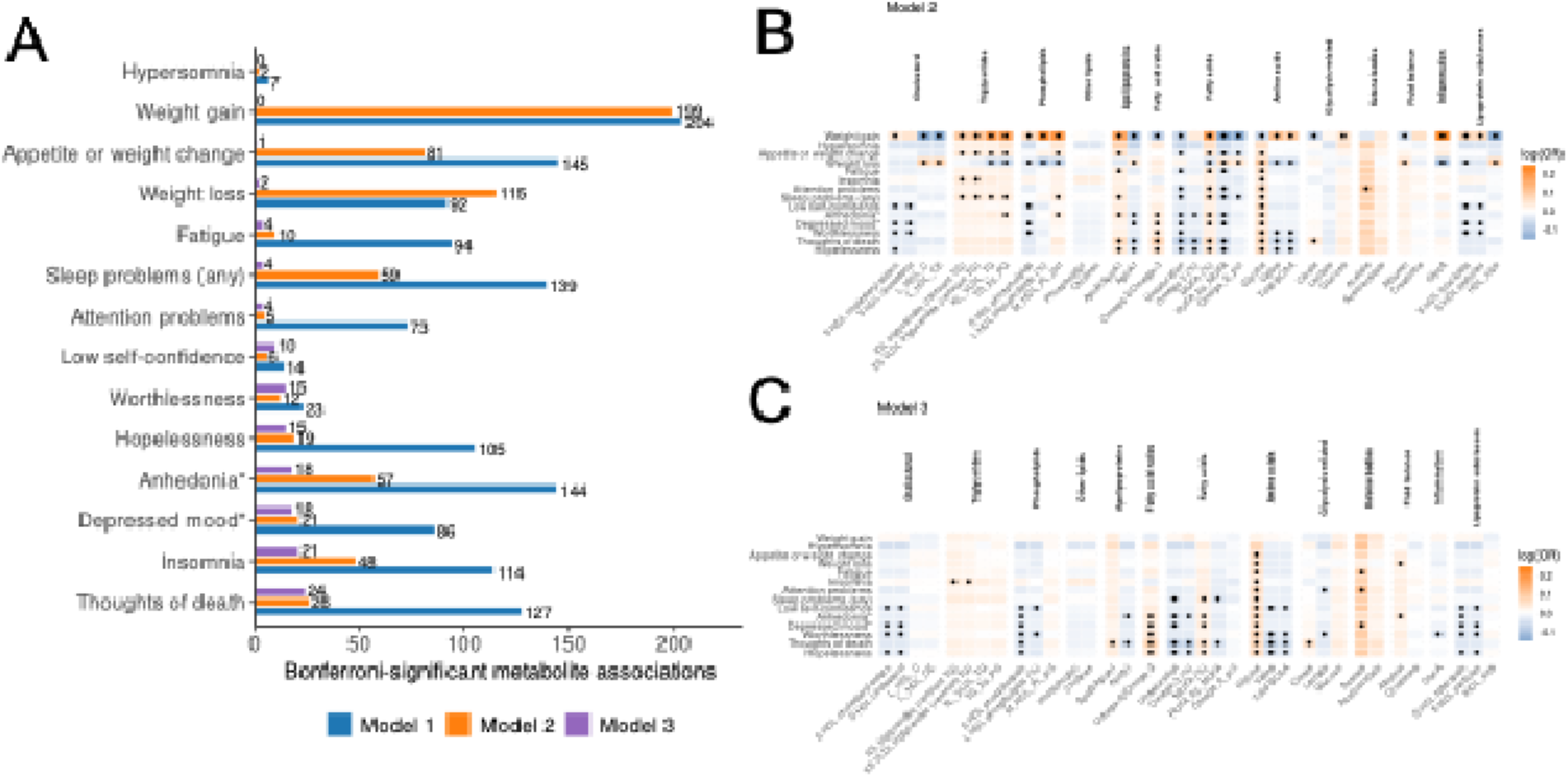
Associations between 14 depressive symptoms and 249 NMR blood metabolites. **(A)** Number of statistically significant associations (Bonferroni-corrected p < 0.05 / (249 × 14)) by symptom and model, ordered by Model 3. Model 1 adjusts for age, sex, and time between sampling and questionnaire; Model 2 additionally adjusts for lifestyle and medication; Model 3 further includes BMI. **(B-C)** Heatmaps of symptom–metabolite associations from Models 2 and 3, respectively; Model 3 additionally includes BMI. Colors indicate log-transformed odds ratios (blue = lower odds, orange = higher odds; white = null), with values capped for comparability; black dots denote Bonferroni-significant associations.

### Attenuation after BMI adjustment

The impact of BMI adjustment on symptom-metabolite associations is illustrated in Figure 2A. The largest shifts from the diagonal are observed for triglyceride-rich lipoproteins, HDL phospholipid measures, fatty-acid composition traits, and GlycA, indicating substantial reduction in effect sizes after BMI adjustment. In contrast, many amino acids and small HDL subclasses cluster closer to the identity line, indicating limited change between models.

**Figure 2.**
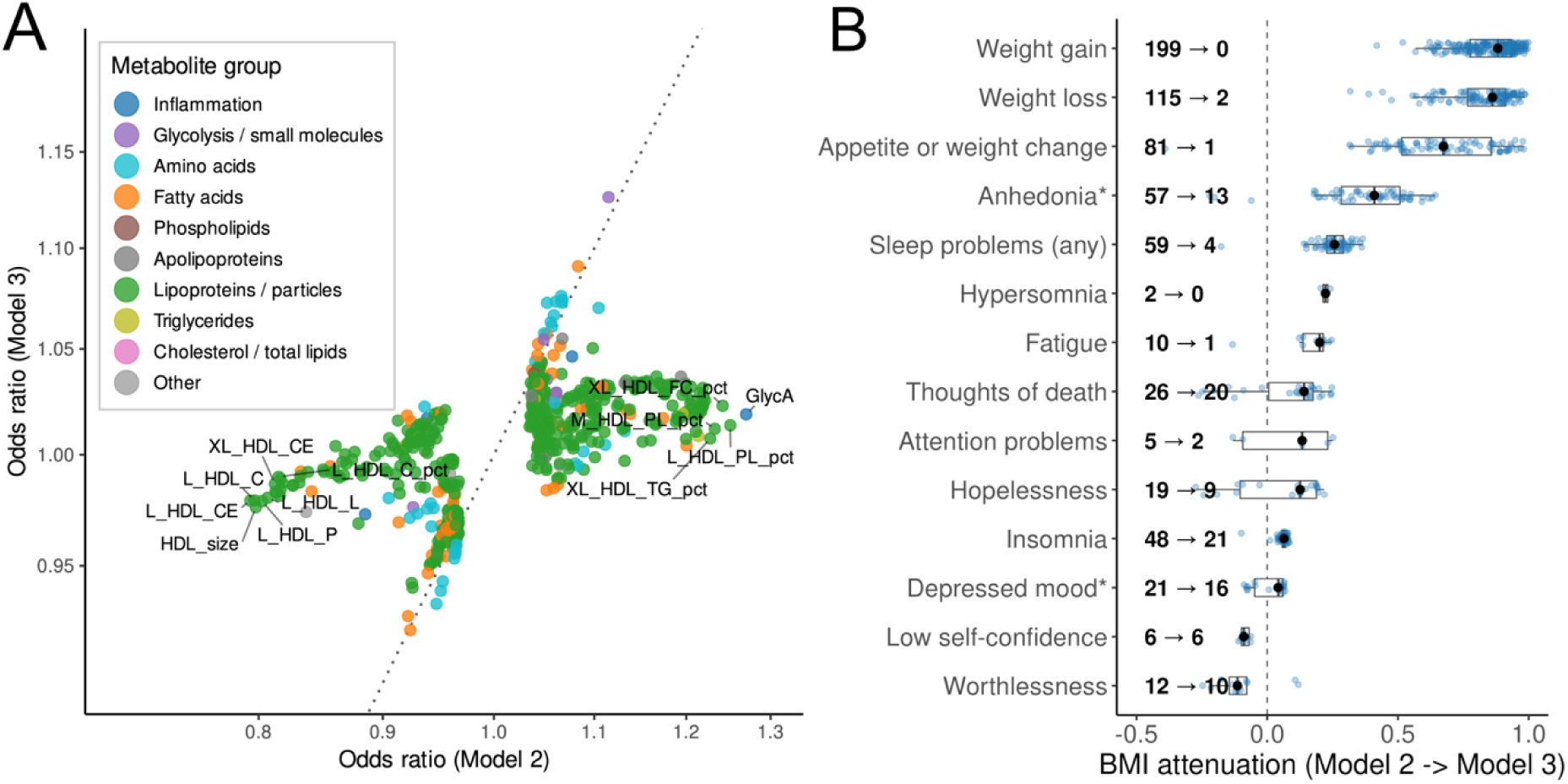
Impact of BMI adjustment on symptom–metabolite associations. **(A)** Comparison of effect estimates between Models 2 and 3. Scatterplot of odds ratios (ORs) for all Bonferroni-significant metabolite–symptom pairs identified in Model 2. The diagonal (x = y) indicates identical estimates; deviation reflects change after BMI adjustment. **(B)** Distribution of BMI attenuation across depressive symptoms. Box plots show attenuation values for all associations significant in Model 2. Points represent individual associations (jittered), and black circles indicate median attenuation per symptom. Symptoms are ordered by median attenuation; the dashed line marks zero attenuation. Text annotations indicate the number of significant associations in Model 2 and those remaining in Model 3.

Symptom-level attenuation patterns are shown in Figure 2B and detailed in Supplementary Table S5. Weight- and appetite-related symptoms showed the strongest BMI sensitivity. For weight gain, none of the 199 Model 2 associations (0.0%) remained Bonferroni-significant in Model 3; for weight loss, 2/115 (1.7%) persisted; and for appetite change, 1/81 (1.2%) remained. Median attenuation coefficients were 0.88 (weight gain), 0.86 (weight loss), and 0.67 (appetite change). Sleep and energy-related symptoms showed intermediate attenuation. For insomnia, 21/48 (43.8%) associations remained significant; for fatigue, 1/10 (10.0%); and for attention problems, 2/5 (40.0%) were retained. Median attenuation coefficients were 0.06, 0.2, and 0.13, respectively.

Among affective symptoms, depressed mood and thoughts of death were comparatively BMI-robust, with 16/21 (76.2%) and 20/26 (76.9%) associations retained, whereas 9/19 (47.4%) hopelessness and 13/57 (22.8%) anhedonia associations persisted (Supplementary Table S5). At the individual metabolite level, attenuation was most pronounced for lipid and inflammatory measures. For example, the GlycA–weight gain association decreased from OR 1.27 (95% CI 1.24–1.30) in Model 2 to OR 1.02 (95% CI 0.99–1.05) in Model 3. Similarly, L-HDL-PL% attenuated from OR 1.25 to 1.01.

A subset of associations showed limited attenuation and remained Bonferroni-significant in Model 3 (Supplementary Table S5). These BMI-robust associations primarily involved amino acids, fatty-acid unsaturation measures, and small HDL particle subclasses. For example, glycine was associated with loss of interest (OR 1.073, 95% CI 1.057–1.089), and acetate with attention difficulties (OR 1.126, 95% CI 1.076–1.179). Higher fatty-acid unsaturation was inversely associated with thoughts of death (OR 0.928, 95% CI 0.910–0.947). Small HDL subclasses (S-HDL-PL, S-HDL-CE, S-HDL-L) showed stable inverse associations across multiple symptoms, whereas total HDL cholesterol did not (Supplementary Table S5).

### BMI interaction analysis

Among the 691 preselected pairs, 211 (30.5%) showed metabolite × BMI interaction after FDR correction (Supplementary Table S6). Interactions were concentrated in weight gain (144) and weight loss (48), followed by anhedonia (13); the remaining significant interactions involved thoughts of death, depressed mood, sleep problems, fatigue, and worthlessness. Interaction ORs ranged from approximately 0.92 to 1.08 per 1-SD metabolite × 1-SD BMI. Representative interactions illustrate both steeper and flatter metabolite–symptom slopes with increasing BMI (Figure 3).

**Figure 3.**
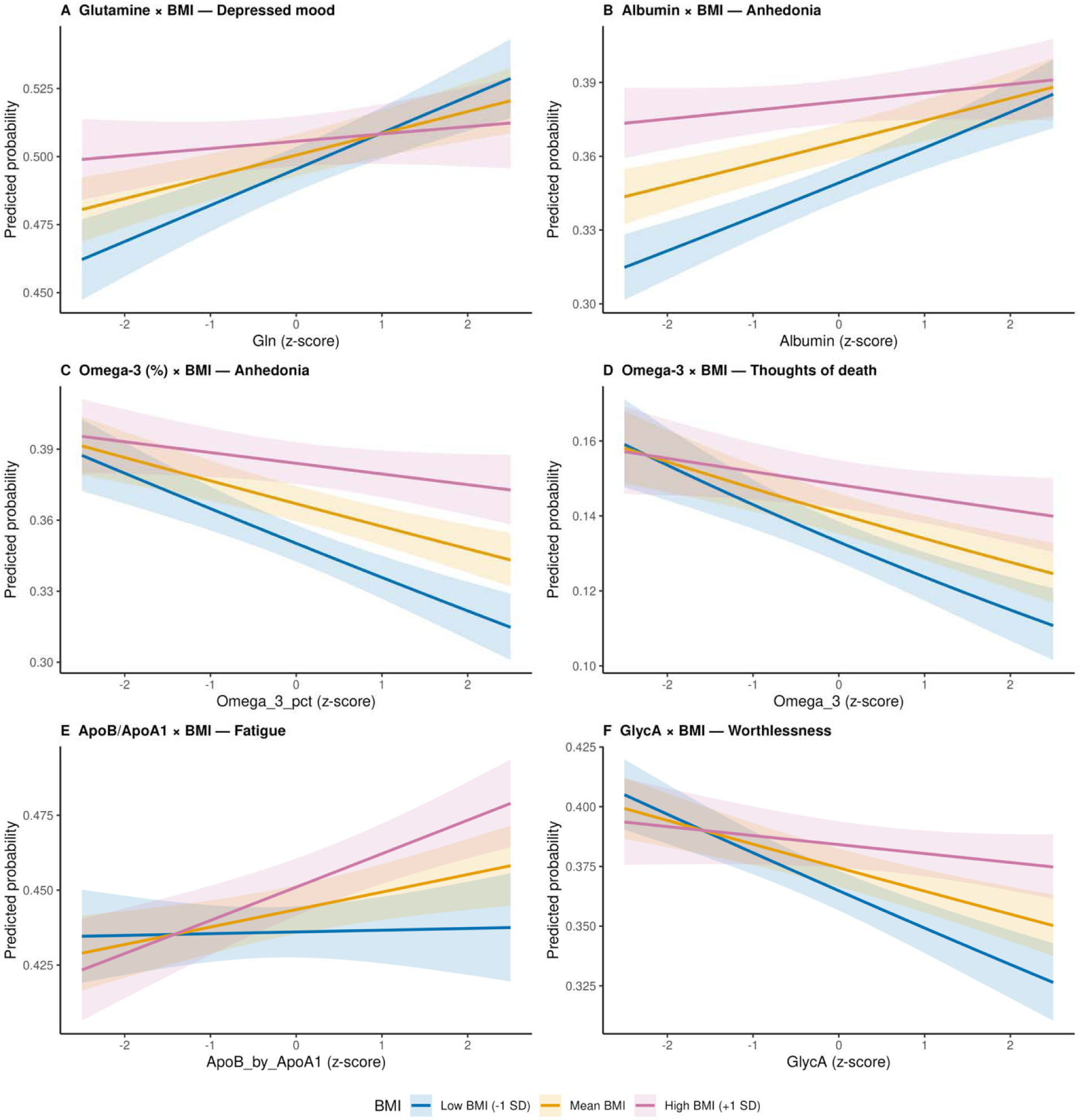
BMI modifies metabolite–symptom associations. **(A–F)** Predicted probabilities of depressive symptoms based on metabolite × BMI interaction terms, shown across standardized metabolite levels (z-scores) at three BMI levels (−1 SD, mean, +1 SD). Models are adjusted for age, sex, time between blood sampling and questionnaire completion, smoking, alcohol use, education, and medication. Shaded bands indicate 95% confidence intervals. Panels show interactions for: (A) glutamine and depressed mood, (B) Albumin and anhedonia, (C) omega-3 fatty acid and anhedonia, (D) omega-3 fatty acids and thoughts of death, (E) ApoB/ApoA1 and fatigue, and (F) GlycA and worthlessness.

### Metabolic context of symptom-metabolite associations

The six SOM-derived profiles showed distinct systemic metabolic characteristics, including differences in BMI, lipoprotein measures, fatty acid composition, and GlycA, despite the profiles being derived exclusively from metabolomic data and independently of psychiatric variables and BMI (Figure 4A). To assess whether metabolite–symptom associations differed across systemic metabolic states, we tested metabolite × SOM-profile interactions among the 211 metabolite–symptom pairs with FDR-significant metabolite × BMI interactions.

**Figure 4.**
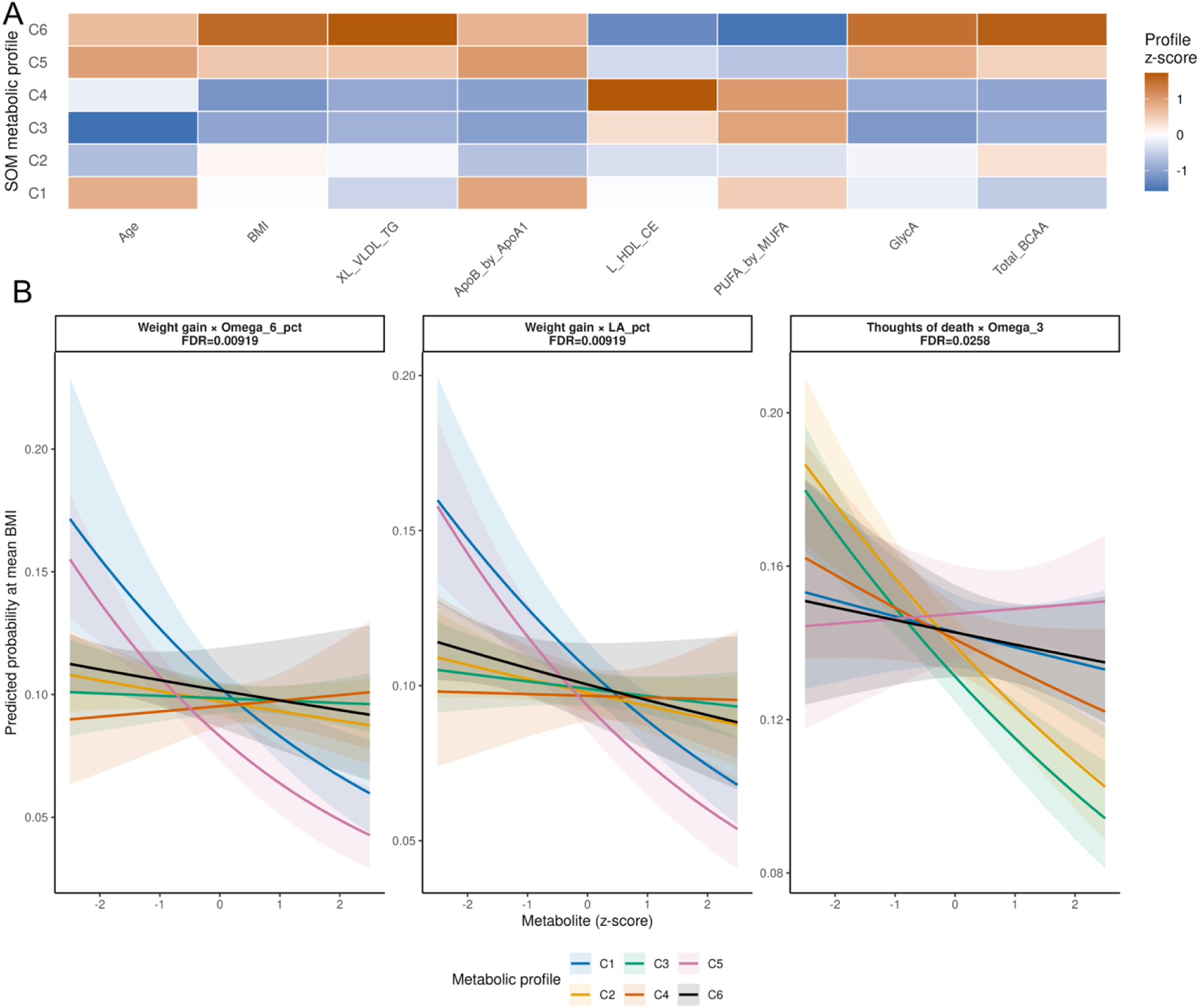
Systemic metabolic differentiation and metabolic profile-dependent metabolite–symptom associations. *(A)* SOM-derived profiles represent distinct systemic metabolic states, differing in age, BMI, lipoprotein measures, fatty acid composition, and GlycA. Profiles were derived exclusively from metabolomic data, independent of psychiatric variables and BMI. ***(B)*** Predicted probabilities across standardized metabolite levels stratified by SOM profile for metabolite–symptom pairs showing significant metabolite × profile interactions after FDR correction. Predictions are shown at mean BMI from models additionally accounting for metabolite × BMI interaction. Shaded areas indicate 95% confidence intervals. FDR values refer to the global metabolite × SOM-profile likelihood ratio test corrected across 211 follow-up tests.

After FDR correction, three metabolite–symptom pairs showed evidence of profile-dependent heterogeneity (FDR < 0.05, Figure 4B, Supplementary Table S9). Weight gain × omega-6 percentage showed the strongest evidence (LRT p = 8.4 × 10 ; FDR = 0.009), with inverse associations most evident in C1 (OR = 0.79, 95% CI 0.69–0.90) and C5 (OR = 0.75, 95% CI 0.67–0.84), while associations were close to null in the other profiles. A similar pattern was observed for weight gain × linoleic acid percentage (LRT p = 8.7 × 10 ; FDR = 0.009), with stronger inverse associations in C1 (OR = 0.83, 95% CI 0.75–0.90) and C5 (OR = 0.79, 95% CI 0.72–0.86).

The third interaction involved thoughts of death × omega-3 fatty acids (LRT p = 3.67 × 10 ; FDR = 0.026). Inverse associations were most pronounced in C2 (OR = 0.87, 95% CI 0.82–0.92) and C3 (OR = 0.86, 95% CI 0.82–0.91), whereas associations were near null in the remaining profiles. Overall, additional profile-dependent heterogeneity was uncommon among metabolite– symptom associations already showing BMI effect modification, with only three of 211 pairs showing significant heterogeneity across SOM profiles after continuous BMI moderation was accounted for.

## Discussion

In this large population-based metabolomic study, circulating metabolic correlates of depressive symptoms varied markedly across symptom domains and BMI-related metabolic context.

Weight- and appetite-related symptoms showed the broadest associations with lipid and inflammatory measures and the strongest attenuation after BMI adjustment, whereas selected affective and cognitive associations were comparatively robust. Formal interaction analyses further showed that BMI modified 30.5% of preselected metabolite–symptom associations. Together, these findings support the view that depression comprises biologically heterogeneous processes and that peripheral metabolic signals are context dependent rather than uniform features of the disorder (5,19,31).

The strongest metabolic associations were observed for weight gain, weight loss, and appetite change and involved triglyceride-rich lipoproteins, fatty-acid measures, and GlycA. These markers are related to hepatic lipid and glucose metabolism, insulin signaling, and systemic inflammation (32–34), consistent with immunometabolic models in which neurovegetative symptoms co-occur with cardiometabolic dysregulation (19,31). Previous metabolomic studies have similarly linked lipid and lipoprotein measures to depressive symptoms (9,35). Our findings extend this work by showing that these associations are concentrated in specific symptoms and are particularly sensitive to BMI-related physiology.

Affective and cognitive symptoms showed fewer associations overall, but several signals persisted after BMI adjustment, including associations involving amino acids, fatty-acid composition, and small HDL subclasses. Altered branched-chain amino-acid concentrations have previously been reported in major depression (36), while amino-acid metabolism and glutamatergic signaling are linked to biological processes implicated in depression (37–39). These findings suggest that not all peripheral metabolic correlates of depressive symptoms are reducible to adiposity-linked variation, although their mechanistic interpretation requires longitudinal or experimental evidence.

BMI is a major determinant of systemic metabolic organization, influencing lipid metabolism, inflammation, and insulin sensitivity (15,40). Its inclusion markedly attenuated many weight-and appetite-related associations, but this effect was not uniform across symptoms. Formal interaction analyses further showed that BMI modified metabolite–symptom relationships rather than acting only as an adjustment factor. Significant interactions were concentrated in weight-related symptoms but were also observed for selected affective, cognitive, and somatic symptoms, consistent with broader adiposity- and inflammation-related metabolic states conditioning associations between circulating metabolites and depressive symptoms (17,18).

Broader systemic metabolic profiles contributed comparatively little additional heterogeneity beyond continuous BMI. Depressive symptom prevalence did not cluster strongly within any single profile, indicating that depression did not correspond to one discrete systemic metabolic state. This differs from recent UK Biobank work that identified three metabolic subtypes by clustering individuals already selected for current depression (41). In our study, profiles were derived in the full population independently of psychiatric phenotype and were used to test effect modification. Among the 211 associations showing BMI effect modification, only three showed additional profile-dependent heterogeneity, involving weight gain with omega-6 and linoleic acid proportions and thoughts of death with omega-3 fatty acids. Thus, among BMI-sensitive associations, broader metabolic state contributed limited additional heterogeneity.

These findings have implications for metabolic biomarker research in psychiatry. Rather than seeking universal metabolic markers of MDD, future studies may gain precision by considering symptom phenotype together with metabolic context. The concentration of immunometabolic signals in neurovegetative symptoms is consistent with work linking metabolic dysregulation to atypical and energy-related depressive features (19,42). However, the present observational findings do not establish treatment targets; instead, they motivate testing whether joint symptom and metabolic stratification improves prediction of prognosis or response to metabolic or anti-inflammatory interventions.

Strengths of this study include the large population-based sample, comprehensive Nightingale NMR profiling, symptom-level phenotyping, and complementary analyses of BMI adjustment, effect modification, and multivariate metabolic context. Several limitations should be considered. Depressive symptoms were assessed retrospectively as lifetime occurrences, whereas metabolites were measured once at recruitment, preventing episode-specific or causal inference. Because secondary symptoms were queried only after endorsement of a core symptom, these outcomes represent population-level lifetime symptom phenotypes; participants without core symptoms were classified as controls. Associations with secondary symptoms may therefore partly reflect differences between individuals with and without core depressive symptoms rather than the secondary symptom in isolation. BMI was assessed at MHoS completion, on average approximately four years after metabolomic sampling; metabolite × BMI interactions therefore describe effect modification by later BMI rather than contemporaneous adiposity. Residual confounding by diet, physical activity, and other health behaviors remains possible, and BMI does not capture body composition or fat distribution. The Nightingale platform provides extensive lipid coverage but more limited representation of other biochemical pathways. The SOM also showed limited local topology preservation (TE = 0.81); accordingly, inference relied on broader metabolic profiles rather than adjacency between individual SOM units. Finally, the predominantly European-ancestry Estonian Biobank population may limit generalizability to more diverse or clinically ascertained populations.

In conclusion, depression-related metabolic associations are strongly symptom specific and frequently conditioned by BMI-related physiology. Future longitudinal and genetically informed studies should determine whether these patterns reflect shared liability, reverse causation, or causal metabolic mechanisms. Accounting jointly for symptom phenotype and metabolic state may improve the reproducibility and interpretation of metabolic biomarkers in depression.

## Acknowledgements

We thank the Estonian Biobank participants and the Estonian Biobank Research Team: Andres Metspalu, Lili Milani, Tõnu Esko, Mait Metspalu, Priit Palta, Nele Taba, Erik Abner, Jaanika Kronberg, Urmo Võsa. HPC Data analysis was carried out in part in the High-Performance Computing Center of University of Tartu.

## Data availability

Individual-level Estonian Biobank data are not publicly available because of participant privacy and data-protection requirements. Access to Estonian Biobank data may be requested through the Estonian Biobank according to its established data-access procedures and is subject to approval of a research proposal and applicable data-use requirements. The present analyses used data release application 6-7/GI/10083 V16.

## Code availability

Analysis code used to generate the primary statistical analyses, supplementary tables and figures is available at https://github.com/skurvits/estbb-depression-metabolomics.

## Author contributions

S.K. performed all analyses, interpreted the results, prepared the figures and tables, and drafted the manuscript. N.T. prepared and normalized the metabolomics data, contributed to study design, modelling and covariate selection, and reviewed the manuscript. L.M., T.H. and K.L. supervised the study and contributed expertise in genomics, metabolomics and psychiatric genetics, respectively, as well as interpretation and manuscript revision. The E.B.R.T. contributed to data generation and biobank resources. All authors reviewed and approved the final manuscript.

## Declaration of AI-assisted technologies

During manuscript preparation, the authors used ChatGPT (OpenAI) to assist with language editing, restructuring of selected passages, and checks of clarity and consistency. The tool was not used to conduct statistical analyses, generate study results, or make scientific conclusions. All AI-assisted text was critically reviewed, revised, and verified by the authors, who take full responsibility for the final manuscript.

## References

1. World Health Organization (2017): Depression and Other Common Mental Disorders: Global Health Estimates. Geneva: World Health Organization.

2. Marx W, Penninx BWJH, Solmi M, Furukawa TA, Firth J, Carvalho AF, Berk M (2023): Major depressive disorder. Nat Rev Dis Primers 9: 44.

3. American Psychiatric Association (2013): Diagnostic and Statistical Manual of Mental Disorders, 5th ed. Washington, DC: American Psychiatric Publishing.

4. World Health Organization (2016): International Statistical Classification of Diseases and Related Health Problems, 10th Revision. Geneva: World Health Organization.

5. Fried EI, Nesse RM (2015): Depression is not a consistent syndrome: An investigation of unique symptom patterns in the STAR*D study. J Affect Disord 172: 96–102.

6. Fried EI (2017): The 52 symptoms of major depression: Lack of content overlap among seven common depression scales. J Affect Disord 208: 191–197.

7. Beijers L, Wardenaar KJ, van Loo HM, Schoevers RA (2019): Data-driven biological subtypes of depression: Systematic review of biological approaches to depression subtyping. Mol Psychiatry 24: 888–900.

8. Malgaroli M, Calderon A, Bonanno GA (2021): Networks of major depressive disorder: A systematic review. Clin Psychol Rev 85: 102000.

9. Bot M, Milaneschi Y, Al-Shehri T, Amin N, Garmaeva S, Onderwater GLJ, et al. (2020): Metabolomics profile in depression: A pooled analysis of 230 metabolic markers in 5283 cases with depression and 10,145 controls. Biol Psychiatry 87: 409–418.

10. Jansen R, Milaneschi Y, Schranner D, Kastenmuller G, Arnold M, Han X, et al. (2024): The metabolome-wide signature of major depressive disorder. Mol Psychiatry 29: 3722–3733.

11. de Kluiver H, Jansen R, Penninx BWJH, Giltay EJ, Schoevers RA, Milaneschi Y (2023): Metabolomics signatures of depression: The role of symptom profiles. Transl Psychiatry 13: 198.

12. Bernhardsen GP, Thomas O, Mäntyselkä P, Niskanen L, Vanhala M, Koponen H, Lehto SM (2024): Metabolites and depressive symptoms: Network- and longitudinal analyses from the Finnish Depression and Metabolic Syndrome in Adults (FDMSA) Study. J Affect Disord 347: 199–209.

13. Alshehri T, Mook-Kanamori DO, Willems van Dijk K, Dinga R, Penninx BWJH, Rosendaal FR, et al. (2023): Metabolomics dissection of depression heterogeneity and related cardiometabolic risk. Psychol Med 53: 248–257.

14. Milaneschi Y, Kappelmann N, Ye Z, Lamers F, Moser S, Jones PB, et al. (2021): Association of inflammation with depression and anxiety: Evidence for symptom-specificity and potential causality from UK Biobank and NESDA cohorts. Mol Psychiatry 26: 7393–7402.

15. Després JP (2012): Body fat distribution and risk of cardiovascular disease: An update. Circulation 126: 1301–1313.

16. Kivimäki M, Jokela M, Hamer M, Geddes J, Ebmeier K, Kumari M, et al. (2011): Examining overweight and obesity as risk factors for common mental disorders using fat mass and obesity-associated (FTO) genotype-instrumented analysis: The Whitehall II Study, 1985-2004. Am J Epidemiol 173: 421–429.

17. Zwiep JC, Milaneschi Y, Giltay EJ, Vinkers CH, Penninx BWJH, Lamers F (2025): Depression with immuno-metabolic dysregulation: Testing pragmatic criteria to stratify patients. Brain Behav Immun 124: 115–122.

18. Capuron L, Lasselin J, Castanon N (2017): Role of adiposity-driven inflammation in depressive morbidity. Neuropsychopharmacology 42: 115–128.

19. Penninx BWJH, Lamers F, Jansen R, Berk M, Khandaker GM, De Picker L, et al. (2025): Immuno-metabolic depression: From concept to implementation. Lancet Reg Health Eur 48: 101166.

20. Insel TR, Cuthbert BN (2015): Brain disorders? Precisely. Science 348: 499–500.

21. Raison CL, Rutherford RE, Woolwine BJ, Shuo C, Schettler P, Drake DF, et al. (2013): A randomized controlled trial of the tumor necrosis factor antagonist infliximab for treatment-resistant depression: The role of baseline inflammatory biomarkers. JAMA Psychiatry 70: 31–41.

22. Savitz J, Figueroa-Hall LK, Teague TK, Yeh HW, Zheng H, Kuplicki R, et al. (2025): Systemic inflammation and anhedonic responses to an inflammatory challenge in adults with major depressive disorder: A randomized controlled trial. Am J Psychiatry 182: 560–568.

23. Nightingale Health Biobank Collaborative Group (2024): Metabolomic and genomic prediction of common diseases in 700,217 participants in three national biobanks. Nat Commun 15: 10092.

24. Ojalo T, Haan E, Kõiv K, Kariis HM, Krebs K, Uusberg H, et al. (2024): Cohort profile update: Mental health online survey in the Estonian Biobank (EstBB MHoS). Int J Epidemiol 53: dyae017.

25. Milani L, Alver M, Laur S, Reisberg S, Haller T, Aasmets O, et al. (2025): The Estonian Biobank’s journey from biobanking to personalized medicine. Nat Commun 16: 3270.

26. Kessler RC, Andrews G, Mroczek D, Ustun B, Wittchen HU (1998): The World Health Organization Composite International Diagnostic Interview Short Form (CIDI-SF). Int J Methods Psychiatr Res 7: 171–185.

27. Ritchie SC, Surendran P, Karthikeyan S, Lambert SA, Bolton T, Pennells L, et al. (2023): Quality control and removal of technical variation of NMR metabolic biomarker data in ∼120,000 UK Biobank participants. Sci Data 10: 64.

28. Meinicke P, Lingner T, Kaever A, Feussner K, Göbel C, Feussner I, et al. (2008): Metabolite-based clustering and visualization of mass spectrometry data using one-dimensional self-organizing maps. Algorithms Mol Biol 3: 9.

29. Wehrens R, Buydens LMC (2007): Self- and super-organizing maps in R: The kohonen package. J Stat Softw 21: 1–19.

30. Wehrens R, Kruisselbrink J (2018): Flexible self-organizing maps in kohonen 3.0. J Stat Softw 87: 1–18.

31. Milaneschi Y, Lamers F, Berk M, Penninx BWJH (2020): Depression heterogeneity and its biological underpinnings: Toward immunometabolic depression. Biol Psychiatry 88: 369–380.

32. Bhat N, Mani A (2023): Dysregulation of lipid and glucose metabolism in nonalcoholic fatty liver disease. Nutrients 15: 2323.

33. Uehara K, Santoleri D, Whitlock AEG, Titchenell PM (2023): Insulin regulation of hepatic lipid homeostasis. Compr Physiol 13: 4785–4809.

34. Connelly MA, Otvos JD, Shalaurova I, Playford MP, Mehta NN (2017): GlycA, a novel biomarker of systemic inflammation and cardiovascular disease risk. J Transl Med 15: 219.

35. Rydin AO, Milaneschi Y, Quax R, Li J, Bosch JA, Schoevers RA, et al. (2023): A network analysis of depressive symptoms and metabolomics. Psychol Med 53: 7385–7394.

36. Baranyi A, Amouzadeh-Ghadikolai O, von Lewinski D, Rothenhäusler HB, Theokas S, Robier C, et al. (2016): Branched-chain amino acids as new biomarkers of major depression—a novel neurobiology of mood disorder. PLoS One 11: e0160542.

37. Xu S, Liu Y, Pu J, Gui S, Zhong X, Tian L, et al. (2020): Chronic stress in a rat model of depression disturbs the glutamine-glutamate-GABA cycle in the striatum, hippocampus, and cerebellum. Neuropsychiatr Dis Treat 16: 557–570.

38. Sanacora G, Treccani G, Popoli M (2012): Towards a glutamate hypothesis of depression: An emerging frontier of neuropsychopharmacology for mood disorders. Neuropharmacology 62: 63–77.

39. Duman RS, Aghajanian GK, Sanacora G, Krystal JH (2016): Synaptic plasticity and depression: New insights from stress and rapid-acting antidepressants. Nat Med 22: 238–249.

40. Mäkinen VP, Kettunen J, Lehtimäki T, Kähönen M, Viikari J, Peltonen M, et al. (2023): Longitudinal metabolomics of increasing body-mass index and waist-hip ratio reveals two dynamic patterns of obesity pandemic. Int J Obes (Lond) 47: 453–462.

41. Ma S, Nie Z, Zhang M, Mei J, Zhou E, Hu Z, et al. (2025): Towards precision psychiatry: Metabolomics identifies three biological subtypes of depression. PLOS Digit Health 4: e0001125.

42. Zwiep JC, Lamers F, Vinkers CH, van der Wee NJA, Penninx BWJH, Nawijn L, Milaneschi Y (2026): Inflammation, metabolic dysregulation, and depression profiles related to anhedonia and atypical, energy-related symptoms. Brain Behav Immun 132: 106240.

