## Supplementary 1 for "Body mass index modifies symptom-specific metabolomic associations with depressive symptoms in the Estonian Biobank"

### **Supplementary Note 1. Construction and evaluation of the metabolomic self-organizing map**

###### Analytical objective

The self-organizing map (SOM) analysis was used as an unsupervised, prototype-based approach to summarize high-dimensional variation across the 249 Nightingale NMR metabolite measures and to derive broader systemic metabolic profiles for downstream analyses. The SOM was trained independently of depressive symptoms, BMI, and other psychiatric phenotypes.

The two-dimensional SOM lattice was used primarily to organize codebook vectors and facilitate visualization of metabolomic variation. Broader metabolic profiles were subsequently defined by hierarchical clustering of the SOM codebook vectors. Because projection of 249-dimensional metabolomic data onto a two-dimensional lattice necessarily entails information loss, downstream statistical inference relied on the derived metabolic profiles rather than on local adjacency between individual SOM units.

###### Input data and preprocessing

The SOM input consisted of the 249 metabolite measures used in the primary analyses. Metabolites had undergone the quality-control and normalization procedures described in the main Methods and were standardized using sex-specific z-scores before SOM construction.

Sex-specific standardization was used because many Nightingale metabolites differ substantially in their distributions between females and males. This reduced the likelihood that the unsupervised profiles would primarily reflect sex-related differences in absolute metabolite concentrations.

Age was not included as an input variable and metabolites were not age-standardized for the SOM analysis. Age-related metabolic variation was therefore retained as part of the naturally occurring metabolic heterogeneity in the cohort.

For SOM training, missing metabolite values were replaced with the metabolite-specific mean. Mean imputation was used to retain participants in the unsupervised analysis given the low overall level of missingness. Because mean imputation can reduce within-variable variance and does not reproduce extreme values, this procedure represents a limitation of the SOM analysis.

###### SOM architecture and training

Candidate SOM grid sizes were explored to identify a resolution that provided sufficient representation of metabolomic heterogeneity without excessive fragmentation of the sample across map units. The final SOM used a **21 × 21 hexagonal lattice,** corresponding to 441 map units.

The SOM was fitted using the kohonen R package. The final analysis used the package's **online learning algorithm**. Training was performed with:

- grid size: 21 × 21 units;
- topology: hexagonal;
- training length: rlen = 20000;
- initial learning rate: 0.05;
- final learning rate: 0.01;
- random seed: 2025;
- keep.data = TRUE.

The learning rate decreased linearly from 0.05 to 0.01 during training. The SOM grid used the default **bubble neighborhood function** implemented by kohonen. The package documentation defines rlen as the number of times the complete dataset is presented to the network during training and specifies online learning as the default mode when mode is not explicitly supplied. The learning-rate parameter alpha is used for online but not batch training. The default neighborhood function for somgrid() is bubble.

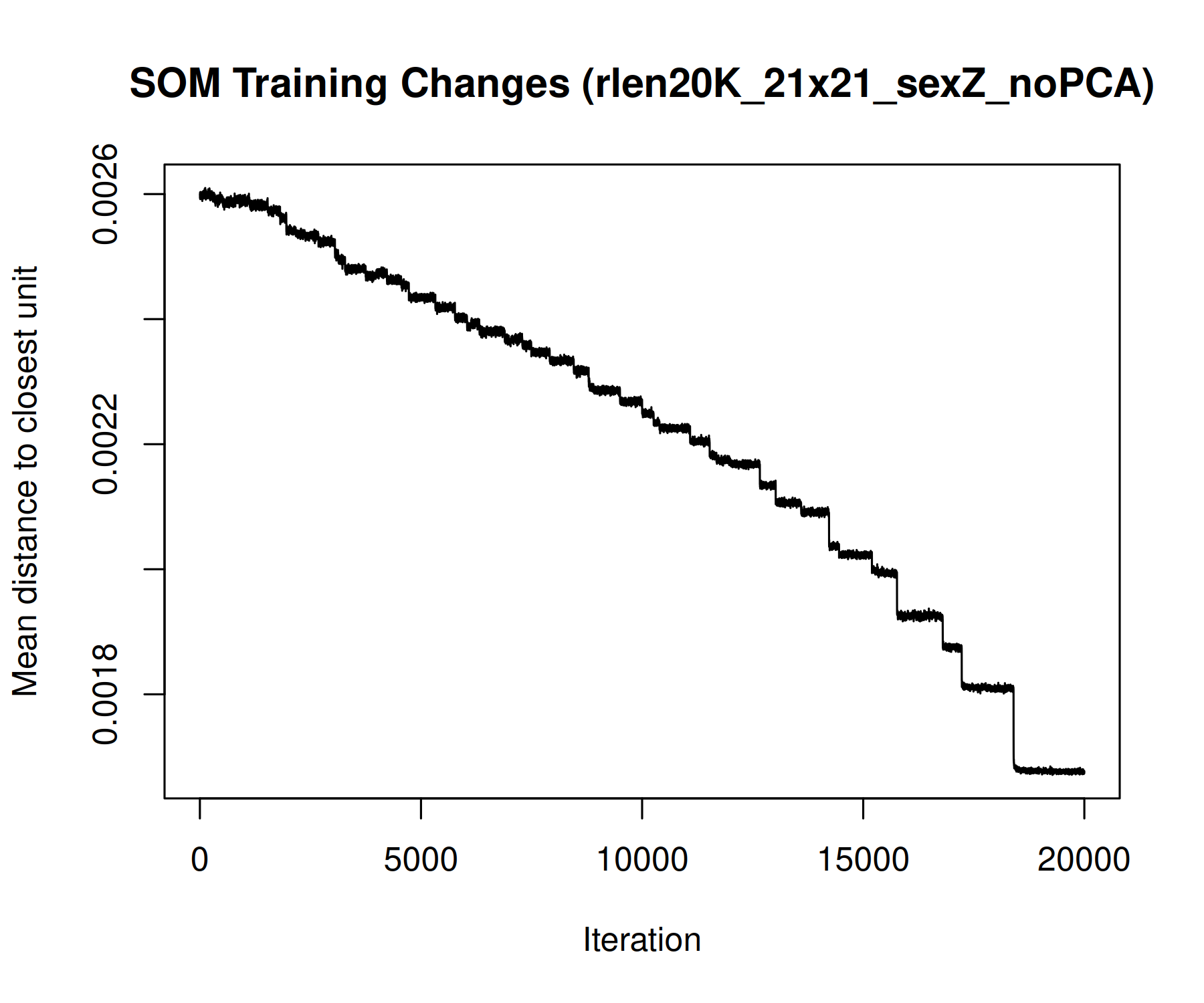

**Figure S1. SOM training changes.**

*Change in mean distance to the closest map unit across SOM training. The plot was used as a descriptive diagnostic of stabilization of the codebook vectors during training.*

#### SOM representation quality

SOM quality was evaluated using complementary metrics and visual diagnostics. No single metric was interpreted as establishing overall validity of the two-dimensional representation.

**Quantization error**

Quantization error (QE) summarizes the distance between each participant's metabolomic profile and the codebook vector of its best-matching unit (BMU). Lower QE indicates closer representation of observations by their assigned SOM prototypes.

The final SOM had a QE of 58.56. Because the absolute magnitude of QE depends on the dimensionality, scaling, and distance metric of the input data, this value was not interpreted against an absolute threshold. QE was used primarily as a comparative diagnostic when evaluating SOM configurations.

**Topographic error**

Topographic error (TE) was calculated as the proportion of observations for which the first- and second-best matching units were not adjacent on the hexagonal lattice. The final SOM had a TE of 0.81, indicating limited preservation of local nearest-neighbor relationships in the two-dimensional representation.

Accordingly, local SOM-unit adjacency was not interpreted as a faithful representation of nearest-neighbor structure in the original 249-dimensional metabolomic space and was not used for statistical inference. The SOM was instead used to obtain prototype vectors that were subsequently aggregated into broader metabolic profiles.

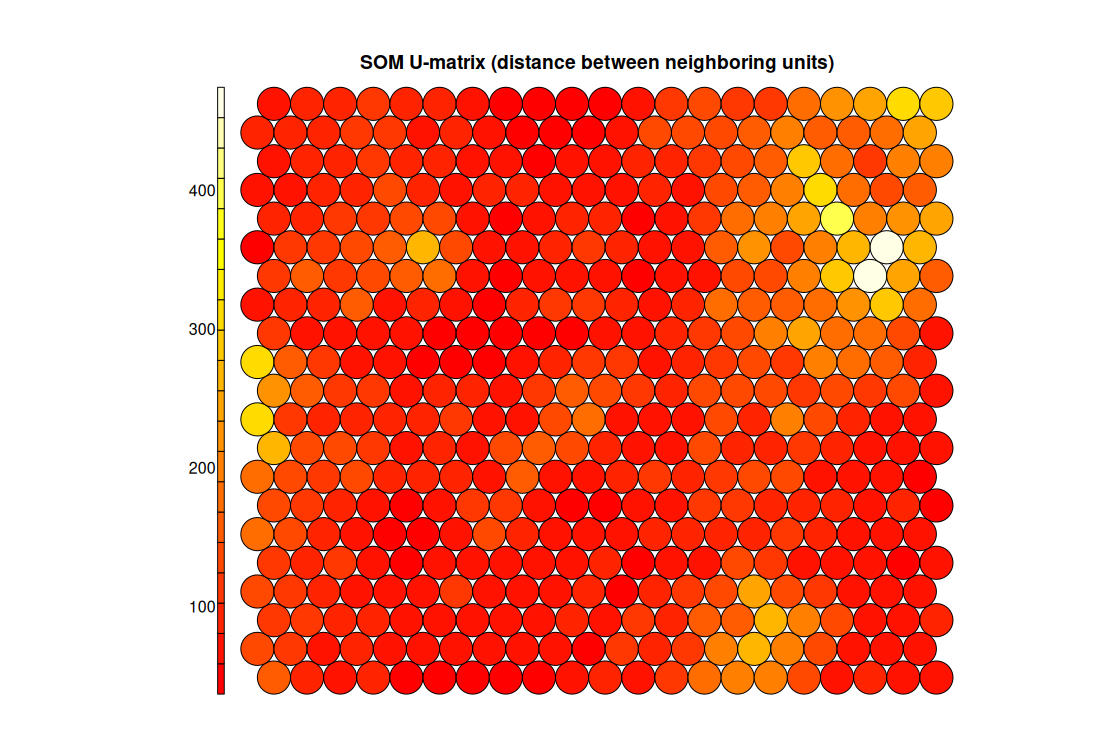

**Figure S2. SOM U-matrix**

*Distances between neighboring SOM codebook vectors. Lower values indicate more similar neighboring prototypes and higher values indicate larger differences between neighboring prototypes. The U-matrix is presented as a descriptive visualization of the fitted SOM and was not used as evidence of preserved local topology.*

###### Visualization of metabolic and phenotypical characteristics

Selected metabolic and participant characteristics were projected onto the trained SOM after model fitting to aid interpretation of the derived metabolic structure. These variables did not contribute to SOM training unless they were among the 249 metabolite inputs.

BMI and depressive symptoms were therefore used only for **post hoc characterization** of the map and derived profiles.

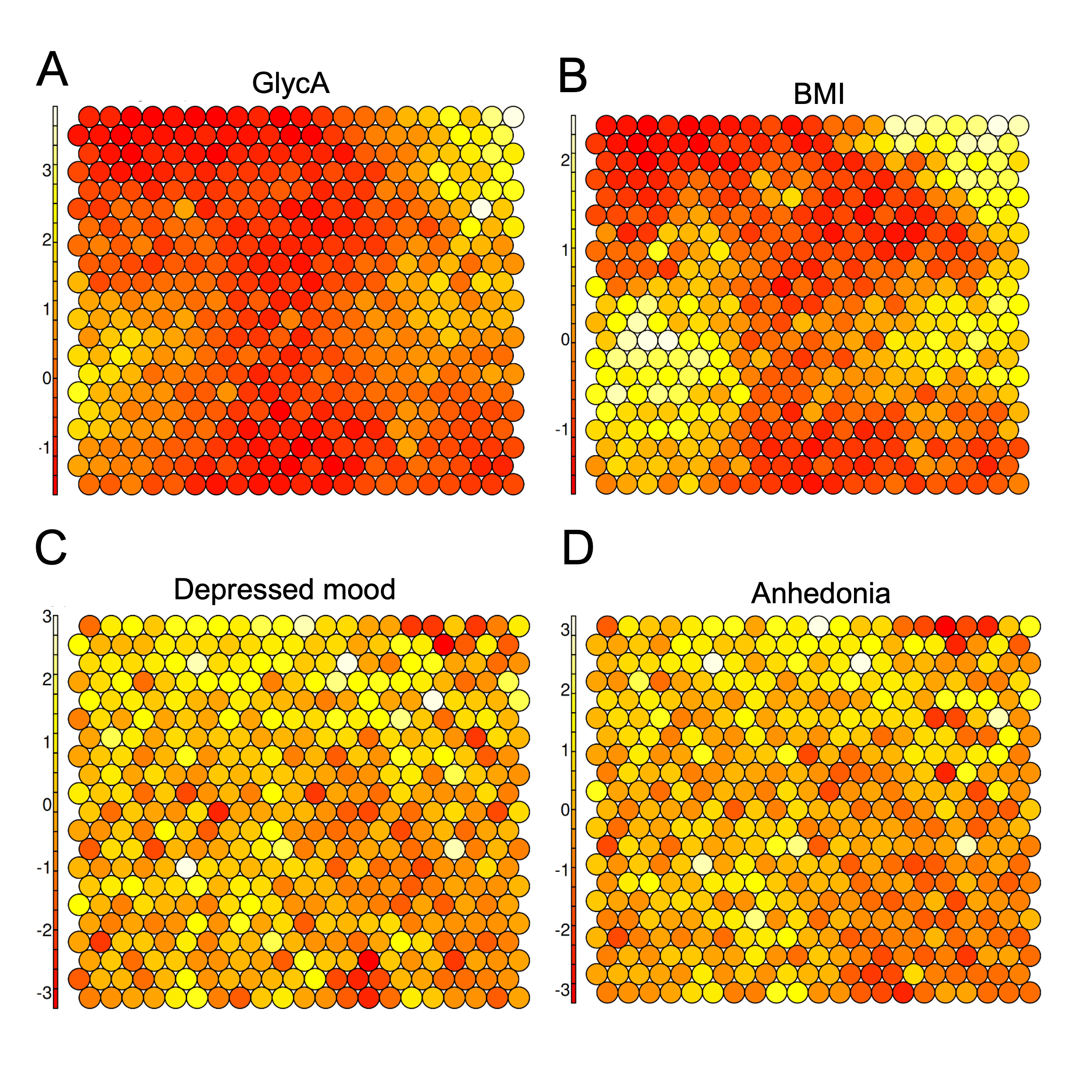

**Figure S3. Distribution of selected metabolic and phenotypic characteristics across the SOM.**
*SOM maps showing mean GlycA levels (A), BMI (B), prevalence of depressed mood (C), and prevalence of anhedonia (D) across map units. The SOM itself was trained exclusively using metabolite measures; BMI and depressive symptoms are overlaid for descriptive interpretation.*

###### Definition of systemic metabolic profiles

Broader metabolic profiles were derived by clustering the SOM codebook vectors rather than clustering participants directly. Euclidean distances between codebook vectors were calculated, followed by hierarchical clustering using the **Ward.D2** method.

Candidate cluster solutions were evaluated over a range of values of K using multiple internal validation indices, including:

- C-index;
- Calinski–Harabasz index;
- Davies–Bouldin index;
- Dunn index;
- silhouette width; and
- Wemmert–Gancarski index.

Higher values indicate more favorable solutions for the Calinski–Harabasz, Dunn, silhouette, and Wemmert–Gancarski indices, whereas lower values are preferable for the C-index and Davies–Bouldin index.

No single value of K was uniformly optimal across all validation metrics. The **six-profile solution (K = 6)** was selected as a pragmatic compromise between internal cluster-validation results, profile size and population coverage, solution complexity, and biological interpretability. We therefore do not interpret K = 6 as a uniquely optimal or biologically definitive number of metabolic states.

After defining the six unit-level clusters, each participant was assigned to the profile containing their best-matching SOM unit.

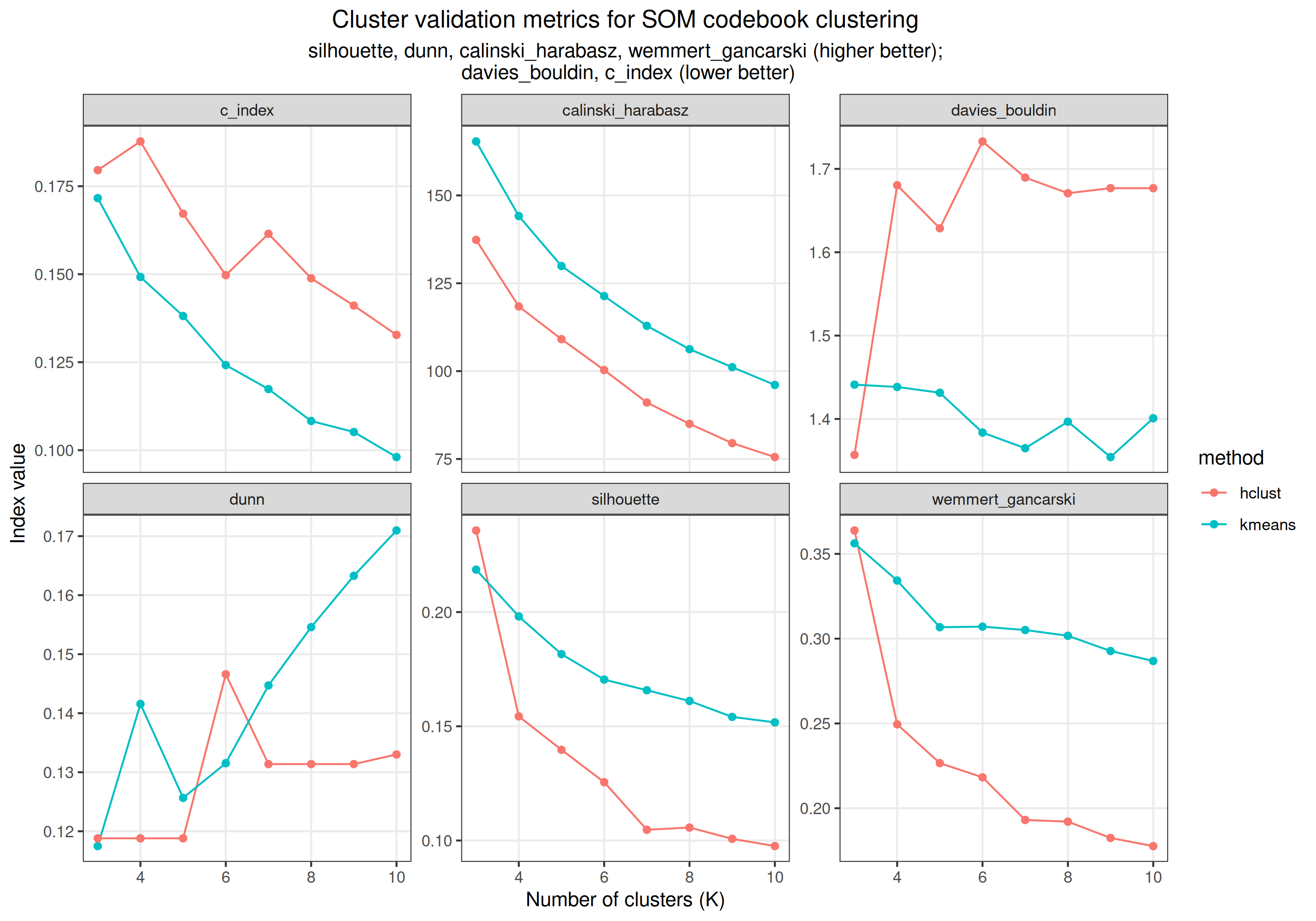

**Figure S4. Internal validation metrics for clustering of SOM codebook vectors.**
*Internal cluster-validation metrics across candidate values of K for Ward.D2 hierarchical clustering. Higher values indicate better solutions for Calinski–Harabasz, Dunn, silhouette, and Wemmert–Gancarski indices; lower values indicate better solutions for the C-index and Davies–Bouldin index. The metrics did not identify a uniformly optimal K across all criteria.*

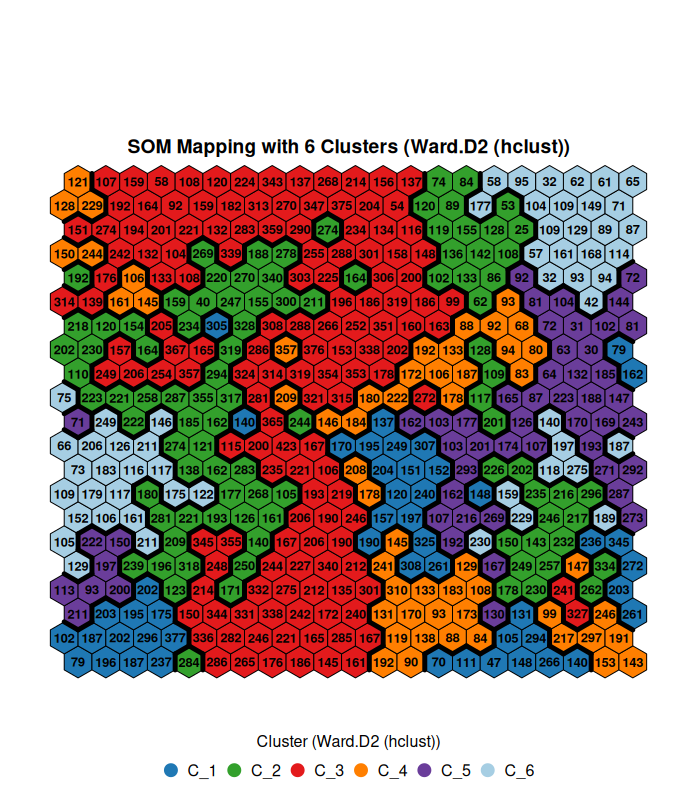

**Figure S5. Six systemic metabolic profiles derived from the SOM.**
*SOM units grouped into six metabolic profiles using Ward.D2 hierarchical clustering of the codebook vectors. Profile boundaries are shown for visualization. Downstream analyses used participant profile membership rather than local relationships between individual SOM units.*

###### Use of SOM profiles in downstream analyses

The SOM profiles were derived independently of BMI and depressive phenotypes and were used as categorical indicators of broader systemic metabolic state.

The primary profile-dependent analysis was restricted to the **211 metabolite–symptom associations that showed FDR-significant metabolite × BMI interaction.** For each of these pairs, a likelihood-ratio test compared a model containing metabolite, BMI, metabolite × BMI interaction, SOM profile, and Model 2 covariates with a model additionally containing metabolite × SOM-profile interaction.

By retaining the continuous metabolite × BMI interaction in both nested models, this analysis tested whether SOM profile explained **additional heterogeneity beyond continuous BMI-dependent effect modification.** Benjamini–Hochberg FDR correction was applied across all 211 profile-interaction tests.

Only three of the 211 tested associations showed FDR-significant additional profile-dependent heterogeneity. Accordingly, the SOM analysis should be interpreted as a secondary analysis of metabolic context rather than as evidence for discrete depression subtypes.

###### Limitation of the SOM analysis

Several features of the SOM analysis should be considered when interpreting the results. First, dimensional reduction from 249 metabolite measures to a two-dimensional lattice entails substantial information loss. Consistent with this, the relatively high topographic error (TE = 0.81) indicates limited preservation of local nearest-neighbor relationships; local map adjacency was therefore not used for inference.

Second, mean imputation of missing metabolite values may reduce variance and does not preserve extreme-value distributions. Third, the six-profile solution represents one useful coarse-grained partition of continuous metabolomic variation rather than a uniquely determined biological classification. Different clustering algorithms, map initializations, or choices of K could yield somewhat different profile boundaries.

Finally, the SOM profiles should not be interpreted as **depression subtypes.** They represent systemic metabolic states derived in the full study population independently of psychiatric phenotypes and were used to evaluate whether broader metabolic context modified individual metabolite–symptom associations.

###### Summary

The SOM analysis provided a prototype-based representation of high-dimensional metabolomic variation from which six broader systemic metabolic profiles were derived. Although preservation of local two-dimensional topology was limited, downstream inference did not depend on local SOM-unit relationships. Instead, the profiles were used as a secondary categorical representation of systemic metabolic context to test for additional heterogeneity in metabolite–symptom associations beyond continuous BMI effect modification.
